# Risk factors for early childhood growth faltering in rural Cambodia

**DOI:** 10.1101/2021.05.20.21257338

**Authors:** Amanda Lai, Irene Velez, Ramya Ambikapathi, Krisna Seng, Oliver Cumming, Joe Brown

## Abstract

**Introduction:** Inadequate nutrition in early life and exposure to sanitation-related enteric pathogens have been linked to poor growth outcomes in children. Despite rapid development in Cambodia, high prevalence of growth faltering and stunting continue to persist. This study aimed to assess nutrition and WASH variables and their association with nutritional status of children under 24 months in rural Cambodia.

**Methods:** We conducted surveys in 491 villages across 55 rural communes in Cambodia in September 2016 to measure associations between child, household, and community-level risk factors for stunting and length-for-age z-score (LAZ). A primary survey measured child-level variables, including anthropometric measures and risk factors for growth faltering and stunting, for 4,036 children under 24 months of age from 3,877 households (approximately 8 households per village). For LAZ, we calculated bivariate and adjusted associations (as mean differences) with 95% confidence intervals using generalised estimating equations (GEEs) to fit linear regression models with robust standard errors. For stunting, we calculated unadjusted and adjusted prevalence ratios (PRs) with 95% confidence intervals using GEEs to fit Poisson regression models with robust standard errors. For all models assessing effects of household-level variables, we used GEEs to account for clustering at the village level.

**Results:** After adjustment for potential confounding, presence of water and soap at a household’s handwashing station was found to be significantly associated (p<0.05) with increased LAZ (adjusted mean difference in LAZ +0.10, 95% CI: 0.03, 0.16), and household use of an improved drinking water source was associated with less stunting in children compared to households that did not use an improved source of drinking water (aPR 0.81, 95% CI: 0.66, 0.98); breastfeeding was associated with a lower LAZ score (−0.16, 95% CI: −0.27, −0.05). No other feeding practices (i.e., dietary diversity, meal frequency, minimum acceptable diet) or sanitation variables (i.e., household’s safe disposal of child stools, household-level sanitation, community-level sanitation) were associated with LAZ scores or stunting in children under 24 months of age. In an age-stratified analysis, children under 12 months of age were longer (LAZ +0.12, 95% CI: 0.02, 0.21) if there was presence of water and soap at the household handwashing station; at the community level, higher prevalence of shared sanitation (percentage of households in a village who report to use shared sanitation facilities) was negatively associated with child length (LAZ - 0.36, 95% CI: −0.66, −0.07).

## Introduction

Childhood growth faltering has been directly linked with adverse outcomes later in life^1^, including poorer school achievement, diminished intellectual functioning, reduced earnings later in life, and lower birthweight for infants born to women who are stunted^2,3^, with the classification of “stunted” defined as having a length-for-age Z-score (LAZ) less than −2 from 2006 WHO International Reference Standard^4^ and “severely stunted” as having a z-score less than −3. Inadequate nutrition has been implicated as a key driver of poor growth outcomes. Interventions that aim to improve child linear growth are typically targeted for children between 6-24 months of age, which is the period critical for cognitive growth and after which is much more difficult to reverse the effects on stunting^5^. On measuring growth outcomes, there is evidence that growth failure at a very young age is strongly linked to shorter adult stature^6^ and puts children at higher risk of death by 24 months of age^7^.

Since growth faltering in children is thought to be primarily attributable to inadequate nutrition, many studies have focused on improving infant and child nutrition ^8–10^ and maternal health^7^ to achieve better growth. However, nutrition behaviours that aim to ensure adequate dietary intake alone have not been successful in eliminating stunting altogether^8^, suggesting the need for additional complementary behaviours that might act synergistically to accelerate progress in countering undernutrition^11^. Enteric infections in early childhood have been shown to impact child growth^12^, primarily via environmental enteric dysfunction^13,14^. Interventions to reduce pathogen exposure, including safe water, effective sanitation, and hygiene (WASH), may therefore play a role in supporting child growth outcomes. These interventions can be directed at both household and community level.

Southeast Asia has seen major reductions in childhood stunting in the last two decades^15^. The prevalence of stunting remains high in Cambodia, however. Cambodia Demographic and Health Survey (CDHS) data from 2014 reported as many as 33% (95% CI: 32%, 34%) of children under five years are stunted and 9% (95% CI: 8.7%, 10%) are severely stunted, defined as having a length-for-age Z-score less than 2 and 3 standard deviations from the WHO reference population^4^; rural populations in Cambodia experience poorer growth outcomes with 36% (95% CI: 34%, 37%) of children under five years stunted and 11% (95% CI: 9.5%, 12%) of children severely stunted^16^. Stunting has been found to be more prevalent among children in rural settings compared to children in urban settings^17,18^, although there is also evidence that poverty – also more prevalent in rural areas – is strongly associated with undernutrition and its risk factors^17^.

The evidence base for sanitation improvements in rural households alone to improve child health is mixed^9,10,19–21^. Increasing sanitation coverage may provide “herd protection” – by reaching a level of sanitation coverage that effectively contains waste to reduce overall exposure to enteric pathogens in a community – and could support improved growth outcomes in children^22–25^. A recent study in Cambodia found community-level open defecation to be associated with decreased length-for-age^26^. Another study of CDHS data (2000-2010) examined risk factors for poor growth outcomes and found a reduction of stunting attributable access to any household sanitation (flush facilities, pit latrines, or composting toilets^27^. Because integrated nutrition and rural sanitation programming are widely being considered as interventions to reduce undernutrition in rural development initiatives ^9,10,21^, this study aims to provide a broad examination of risk factors for undernutrition that focus on child feeding practices and specific household and community-scale WASH measures common in rural Cambodia. Several recent trials^9,10,19,20,28–31^ have sought to measure effects of WASH interventions on growth outcomes in children under two years of age. We examined associations between concurrent WASH and nutritional variables and growth status in children under 24 months in rural Cambodia.

## Methods

### Study and survey design

We measured associations between key WASH and nutrition practices on child linear growth in rural households and villages in three provinces of Cambodia. We conducted this cross-sectional study in 491 villages spanning 55 rural communes of Pursat, Siem Reap, and Battambang provinces in September 2016. Each survey was completed in approximately 30 minutes, and all surveys were completed within a five-week period. Communes were eligible if two key criteria were met: at least 30% of the population lived below the poverty line according to the 2011 Cambodia Ministry of Planning’s Commune Database; and latrine subsidies were not in place, which were both associated with potential short-term changes in sanitation coverage.

We estimated sample size to allow for hypothesis testing in future intervention studies. Using a baseline mean LAZ of −1.64 with a standard deviation of 1.29 from the 2014 CDHS dataset^16^, we estimated this study had 80% power (beta) to detect a minimum detectable effect size (MDES) of 0.18 in length-for-age Z-score at 95% significance (alpha=0.05)^9,19,21^. We used an intra-cluster coefficient of 0.01 using the Cambodia Helping Address Rural Vulnerabilities and Ecosystem Stability (HARVEST) dataset. Complete sample size calculations are provided in the Supplemental Information.

### Study variables

The two primary outcomes were length-for-age Z-score (LAZ, continuous scale) and stunting (dichotomised, defined as LAZ less than −2 standard deviations from the 2006 WHO International Reference Standard^4^) at the individual child-level and at the village-level, expressed as a mean value. Length measurement procedures were performed following Food and Nutrition Technical Assistance (FANTA) guidelines (Supplemental Information). Recumbent lengths were taken per FANTA guidelines, which suggest a recumbent length measurement for children 0-24 months. All anthropometric measurement was performed in duplicate by trained enumerators, and if values differed by >1.0 cm, a third was taken or until successive measurements were <1.0 cm in difference. Final length and weight measurements for z-score calculations were made by taking the mean of the two measurements within the error threshold of 1.0 cm^32^.

The conceptual framework underpinning this analysis is derived from previous literature^12,26,27^ and includes a range of nutrition, water, sanitation, and hygiene variables which could plausibly influence child growth. Child-level nutrition variables included breastfeeding (dichotomous, based on whether child was breastfed yesterday), dietary diversity (dichotomous, based on whether the recommended minimum of four out of seven of food groups was consumed in the previous 24 hours), meal frequency (dichotomous, based on whether the recommended minimum was met), and minimum acceptable diet (dichotomous, based on whether minimum dietary diversity and minimum meal frequencies were met). The household-level water variable included access to an improved drinking water source (dichotomous). The household-level hygiene variable included availability of water and soap at a handwashing station (dichotomous).

Sanitation variables were measured at the household and community level. Household sanitation variables included practice of open defecation (dichotomous), use of a shared sanitation facility (dichotomous), access to an improved sanitation facility (dichotomous), and proper disposal of child stool (dichotomous). Community-level sanitation variables were the same as household-level variables, calculated using village-level means with post-stratification weights (described above).

### Statistical methods

Primary analysis to identify potential risk factors included modelling effects of child-level, household-level, and community-level WASH variables on child-level undernutrition outcomes. For LAZ, we calculated bivariate and adjusted associations (as mean differences) with 95% confidence intervals using generalised estimating equations (GEEs) to fit linear regression models with robust standard errors^33^. For stunting, we calculated unadjusted and adjusted prevalence ratios (PRs) with 95% confidence intervals using GEEs to fit Poisson regression models with robust standard errors^34^. All models assessing effects of household-level variables were adjusted for village level clustering. To test for presence of multicollinearity between covariates, we calculated variance inflation factors (VIFs). All covariates chosen had VIF<5, suggesting no detectable presence of multicollinearity^35^.

Covariates were considered as potential confounders using a “common cause” approach^36^ and on the basis of the conceptual framework describing proposed child feeding practices and WASH variables affecting child nutritional status^12^. In adjusted analyses, we included the following covariates, identified *a priori*: child sex (dichotomous), child age (continuous, in months), child birthweight (continuous, in kilograms), child illness (dichotomous, based on whether caregiver reported any diarrhoea, bloody stool, vomiting, fever, or abdominal pain in the previous week), maternal age (continuous, in years), maternal education (dichotomous, based on whether mother attended primary school or higher), household size (continuous, number of household members), and household wealth index quintile (ordinal).

We performed a complementary analysis to better understand the effects of community-level WASH variables. We used mixed effects regression to model the effects of community-level WASH on LAZ and prevalence stunting, with villages as a fixed effect. GEEs were not used because clustering may have attenuated community-level effects.

## Results

For child-level variables, 4,036 children under 24 months of age from 3,877 households (approximately 8 households per village) were surveyed and had anthropometric measures taken. For some child-level nutrition variables specifically, 2,957 children between 6-23 months of age had dietary diversity scores and meal frequencies measured. For village-level WASH variables, a total of 5,341 households, (approximately 11 households per village) were surveyed.

Table 1 summarises results from the primary survey which captures household, demographic, and WASH characteristics of households with children under 2 years of age. Households had an average size of 5 members with 2-3 children from 2-18 years of age and 1 child below 2 years of age. Most households had a finished floor (95%) and mobile phone (86%), but only 50% had electricity. The mean maternal age was 29.4 year, and most mothers (84%) had attended primary school.

**TABLE 1:** CHILD, HOUSEHOLD, WATER, SANITATION, AND HYGIENE CHARACTERISTICS OF HOUSEHOLDS WITH CHILDREN <24M.

| **HH with children** |  |  |  |
| --- | --- | --- | --- |
|  | N | % or mean | SD |
| **Child characteristics** |  |  |  |
| Child age (months) | 4,064 | 11.1 | 6.6 |
| Male | 4,082 | 0.52 | 0.50 |
| Child birthweight (kg) | 4,033 | 3.07 | 0.46 |
| Currently breastfed (all children) | 3,979 | 77% | 42% |
| Currently breastfed (children 0-6 months) | 1,114 | 98% | 15% |
| Currently breastfed (children 6-12 months) | 1,155 | 91% | 28% |
| Currently breastfed (children 12-18 months) | 943 | 72% | 45% |
| Currently breastfed (children 18-24 months) | 767 | 31% | 46% |
| Solid foods introduced (children 6-8 months) | 521 | 88% | 32% |
| Ever breastfed | 4,082 | 98% | 14% |
| Length for age Z score (LAZ) | 3,984 | -0.96 | 1.16 |
| Stunted | 3,984 | 16% | 37% |
| Caregiver-reported diarrhoea (7-day recall) | 4,082 | 25% | 43% |
| Caregiver-reported diarrhoea (14-day recall) | 4,082 | 7% | 26% |
| Blood in stool (7-day recall) | 4,082 | 2% | 13% |
| Vomit (7-day recall) | 4,082 | 8% | 27% |
| Fever (7-day recall) | 4,082 | 20% | 40% |
| Abdominal pain (7-day recall) | 4,082 | 18% | 39% |
| Any illness | 4,082 | 42% | 49% |
| Minimum dietary diversity met (children >6mon) | 2,957 | 36% | 48% |
| Minimum meal frequency met | 4,082 | 55% | 50% |
| Minimum acceptable diet met (children >6mon) | 2,957 | 37% | 1% |
| **Household characteristics** |  |  |  |
| Household size | 4,082 | 5.5 | 2.2 |
| Number of children in HH (2-18y) | 4,082 | 2.5 | 1.4 |
| Number of children in HH (<24m) | 4,082 | 1.1 | 0.3 |
| Has electricity | 4,082 | 50% | 50% |
| Owns a mobile phone | 4,082 | 85% | 36% |
| Has a finished floor [1] | 4,081 | 95% | 22% |
| Primary caregiver has attended primary school | 4,080 | 84% | 36% |
| Maternal age (years) | 4,066 | 29.4 | 9.1 |
| Improved drinking water source [2] | 4,072 | 85% | 36% |
| Water source on site | 4,082 | 78% | 41% |
| Water source is <5 min, roundtrip | 893 | 13% | 96% |
| Minutes to fetch water, roundtrip | 893 | 17.2 | 23.6 |
| Presence of water at handwashing station | 4,076 | 94% | 24% |
| Presence of soap at handwashing station | 4,076 | 59% | 49% |
| Presence of water and soap at handwashing station | 4,076 | 56% | 50% |
| Had any sanitation facility | 4,075 | 65% | 48% |
| Had improved sanitation facility [3] | 4,082 | 40% | 49% |
| Open defecation | 4,075 | 35% | 48% |
| Used shared toilet | 4,082 | 25% | 43% |
| Child stools properly disposed of [4] | 3,068 | 86% | 35% |
| [1] Finished floor defined as floor made of wood plans, palm/bamboo, parquet or polished wood, vinyl or asphalt strips, ceramic tiles, cement tiles, or cement. Floor materials were classified by enumerator observation. [2] Improved sources of drinking water include: piped water into dwelling/yard/plot, public tap or standpipe, tube well or borehole, protected dug well, protected spring, bottled water, and rainwater. [3] Improved sanitation facilities include: flush/pour flush toilet to a piped sewer system, septic tank or pit latrine, a ventilated improved pit latrine, a pit latrine with slab, and a composting toilet. [4] Proper disposal of children faeces consist of putting or rinsing stool into a sanitation facility or burying it; unsafe disposal of children faeces includes putting or rinsing stool into a drain or ditch, throwing it into garbage or leaving it in the open. |  |  |  |

The average age of children enrolled was 11 months, with approximately 57% (2270/3988) younger than 12 months and 43% (1718/3988) between 12-24 months old. Slightly less than half (47.8 percent) of the children were girls, and the average birth weight was 3.1 kilograms. High prevalence of breastfeeding observed among young children 0-12 months old (94% of children 0-12 months old and 53% of children 12-24 months old). The mean LAZ for all children was - 0.96 (SD 1.16), with older children (12-24 months) having worse growth outcomes (LAZ −1.32, SD 1.16) than younger children (0-12 months, LAZ −0.69, SD 1.06). Similarly, older children (12-24 months) had higher stunting levels (24%, SD 30%) than younger children (0-12 months, 10%, SD 42%). Caregivers reported diarrhoea with a 7-day recall in 25% of children and with a 14-day recall in 7% of children.

Fifty-five percent of all children consumed the recommended minimum frequency of meals^37^, while only of 36% of children over 6 months consumed the recommended minimum dietary diversity. Most households surveyed had an improved drinking water source and water source on site (85% and 78%, respectively), although the survey took place during the rainy season (May – October) so most households collected rainwater for drinking. Most households (94%) also had water at their home’s handwashing station, but only 59% of homes had soap. Sixty-five percent of households had access to any sanitation facility (including 25% with shared facilities), while only 40% of households had access to an improved sanitation facility. Although most of the pour/flush systems were recorded as improved systems that discharged into septic tanks or pit latrines (1971/1976 of pour/flush facilities), there was no record of how wastewater and sludges were managed, so we are unable to determine whether these facilities are safely managed per JMP classification scheme^38^. Most households (86%) properly disposed of child stools by burying stools (46%).

Table 2 summarises results from the secondary survey which captures community WASH practices irrespective of children in the household. Compared to households that had children (Table 1), the community overall had less access to an improved drinking water source (72% vs 85%) but more access to an improved sanitation facility (46% vs 40%) and lower prevalence of open defecation practices (31% vs 35%). The community overall used shared toilets less frequently compared to households with children (10% vs 25%) and practiced safe methods of disposing children’s stools more frequently than households with children (93% vs 86%); methods of stool disposal were qualified as “safe” if the child’s faeces was put into any toilet or latrine^39^. Overall, households with children appear to have poorer sanitation practices than the overall community.

**TABLE 2:** COMMUNITY WASH VARIABLES, CALCULATED USING POST-STRATIFICATION WEIGHTS.

| **Community WASH variables** | **N** | **% or mean** | **SD** |
| --- | --- | --- | --- |
| Had improved sanitation facility [1] | 5,341 | 46% | 31% |
| Open defecation | 5,341 | 31% | 30% |
| Used shared toilet | 5,341 | 10% | 16% |
| Child stools properly disposed of [2] | 5,321 | 93% | 16% |
| [1] Improved sanitation facilities include: flush/pour flush toilet to a piped sewer system, septic tank or pit latrine, a ventilated improved pit latrine, a pit latrine with slab, and a composting toilet. [2] Proper disposal of children faeces consist of putting or rinsing stool into a sanitation facility or burying it; unsafe disposal of children faeces includes putting or rinsing stool into a drain or ditch, throwing it into garbage or leaving it in the open. |  |  |  |

Table 3 summarises unadjusted and adjusted LAZ mean differences and the nutrition and WASH variables of interest. At the child level, unadjusted analyses found breastfeeding practices to be positively associated with growth (LAZ +0.40, 95% CI: 0.30, 0.51). However, these associations were attenuated in the adjusted analysis and were found to be negatively associated with height (LAZ −0.16, 95% CI: −0.27, −0.05). At the household level, unadjusted analyses show that presence of water and soap at handwashing station (LAZ +0.11, 95% CI: 0.03, 0.19) and improved sanitation facility (LAZ +0.16, 95% CI: 0.07, 0.25) to be positive associated with growth compared to those whose families practiced open defecation. Children whose household did not report practicing safe disposal of child stools were shorter than those whose households properly disposed of stools (LAZ −0.15, 95% CI: −0.27, −0.02). In the adjusted analysis, only the presence of water and soap at handwashing stations was associated with taller children (LAZ +0.10, 95% CI: 0.03, 0.16). At the community level, we found no significant associations between WASH variables and child growth in the unadjusted analysis.

**TABLE 3:** LINEAR REGRESSION COEFFICIENT FOR ASSOCIATION BETWEEN NUTRITION AND WASH VARIABLES AND LINEAR GROWTH.

|  | N | Unadjusted effect size | N | Adjusted effect size |
| --- | --- | --- | --- | --- |
| **Child-level variables** |  |  |  |  |
| Currently breastfed (a) | 3449 | 0.40 (0.30, 0.51) | 3709 | -0.16 (-0.27, -0.05) |
| Minimum dietary diversity met (a,c) | 2432 | 0.01 (-0.08, 0.10) | 2421 | 0.05 (-0.03, 0.14) |
| Minimum meal frequency met (a,c) | 2432 | 0.05 (-0.07, 0.17) | 2421 | -0.01 (-0.13, 0.10) |
| Minimum acceptable diet met (a,c) | 2432 | -0.01 (-0.05, 0.02) | 2421 | -0.03 (-0.06, 0.02) |
| **Household-level variables** |  |  |  |  |
| Improved drinking water source [1] (a) | 3481 | 0.05 (-0.06, 0.16) | 3767 | 0.04 (-0.06, 0.13) |
| Presence of water and soap at handwashing (a) | 3483 | 0.11 (0.03, 0.19) | 3771 | 0.10 (0.03, 0.16) |
| Safe disposal of child stool [3] (a) | 2601 | -0.15 (-0.27, -0.02) | 2843 | 0.05 (-0.07, 0.16) |
| Sanitation facility (a) | 3483 |  | 3769 |  |
| Improved [2] |  | 0.16 (0.07, 0.25) |  | 0.05 (-0.03, 0.14) |
| Shared |  | 0.08 (-0.03, 0.20) |  | -0.01 (-0.13, 0.10) |
| None (open defecation) |  | ref |  | ref |
| **Community-level variables** |  |  |  |  |
| Safe disposal of child stool (village-level) [3] (b) | 3475 | 0.04 (-0.19, 0.27) | 3474 | -0.01 (-0.23, 0.20) |
| Improved sanitation facility (village-level) [2] (b) | 3489 | 0.10 (-0.02, 0.23) | 3488 | 0.07 (-0.06, 0.19) |
| Shared sanitation facility (village-level) (b) | 3489 | -0.11 (-0.34, 0.12) | 3488 | -0.19 (-0.42, 0.03) |
| OD (village-level) (b) | 3489 | -0.08 (-0.21, 0.05) | 3488 | -0.03 (-0.16, 0.10) |
| (a) Adjusted for child gender, child age, child illness, maternal age, maternal education, household size, and household wealth index quintile; clustered by village. (b) Adjusted for village-level covariates: % male, mean child age, % with illness, % breastfed, and mean household wealth index quintile. (c) only children >6 months included, per WHO minimum recommended dietary diversity and meal frequencies. [1] Improved sources of drinking water include: piped water into dwelling/yard/plot, public tap or standpipe, tubewell or borehole, protected dug well, protected spring, bottled water, and rainwater. [2] Improved sanitation facilities include: flush/pour flush toilet to a piped sewer system, septic tank or pit latrine, a ventilated improved pit latrine, a pit latrine with slab, and a composting toilet. [3] Safe disposal of children faeces consist of putting or rinsing stool into any sanitation facility; unsafe disposal of children faeces includes putting or rinsing stool into a drain or ditch, throwing it into garbage, burying or leaving it in the open. |  |  |  |  |

We performed a stratified risk factor analysis using age strata of 0-12 months and >12-24 months of age to assess age-associated effects on outcomes, since children under 12 months of age are less mobile and may experience different environmental exposures compared with older children in our cohort. Notably, we found that children under 12 months were longer (LAZ +0.12, 95% CI: 0.02, 0.21) if there was presence of water and soap at the household handwashing station in adjusted analyses (S6), and at the community level we found higher prevalence of shared sanitation (percentage of households in a village who report to use shared sanitation facilities) to be negatively associated with child length (LAZ −0.36, 95% CI: −0.66, −0.07). We found no statistically meaningful associations between other WASH variables and child growth for children >12-24 months of age, after adjusting for all other covariates.

Table 4 summarises unadjusted and adjusted associations between stunting and the nutrition and WASH variables of interest. At the child level, the unadjusted analyses found breastfeeding to be negatively associated with stunting (PR 0.56, 95% CI: 0.47, 0.65). However, this association was attenuated in the adjusted analysis. At the household level, our unadjusted analyses found an improved drinking water source to be negatively associated with stunting (PR 0.80, 95% CI: 0.65-0.97), as well as children in households with access to an improved sanitation facility compared to those who practiced open defecation (PR 0.74, 95% CI: 0.62, 0.88). Many of these associations attenuated in the adjusted analysis, and after adjusting for covariates, we only found a household’s access to an improved drinking water source to be negatively associated with stunting (aPR 0.81, 95% CI: 0.66, 0.98). At the community level, none of the variables assessed were significantly associated with stunting, neither in our unadjusted nor in our adjusted analyses. In our complementary analysis, we assessed the impact of village-level associations by evaluating village-level outcomes, we found no statistically significant association between any nutrition or WASH variables and growth faltering or stunting (see S4 and S5).

**TABLE 4:** PREVALENCE RATIOS FOR ASSOCIATION BETWEEN NUTRITION AND WASH VARIABLES AND STUNTINGs.

|  | N | PR (95% CI) | N | Adjusted PR (95% CI) |
| --- | --- | --- | --- | --- |
| **Child-level variables** |  |  |  |  |
| Currently breastfed (a) | 3449 | 0.56 (0.47, 0.65) | 3437 | 1.03 (0.85, 1.24) |
| Minimum dietary diversity met (a,c) | 2432 | 0.91 (0.77, 1.07) | 2421 | 0.87 (0.74, 1.02) |
| Minimum meal frequency met (a,c) | 2432 | 0.97 (0.79, 1.20) | 2421 | 1.09 (0.89, 1.33) |
| Minimum acceptable diet met (a,c) | 2432 | 0.93 (0.79, 1.09) | 2421 | 0.89 (0.76, 1.04) |
| **Household-level variables** |  |  |  |  |
| Improved drinking water source [1] (a) | 3481 | 0.80 (0.65, 0.97) | 3468 | 0.81 (0.66, 0.98) |
| Presence of water and soap at handwashing (a) | 3483 | 0.92 (0.79, 1.08) | 3471 | 0.95 (0.82, 1.10) |
| Safe disposal of child stool [3] (a) | 2601 | 1.06 (0.82, 1.37) | 2589 | 0.81 (0.63, 1.04) |
| Sanitation facility (a) | 3483 |  | 3470 |  |
| Improved [2] |  | 0.74 (0.62, 0.88) |  | 0.87 (0.74, 1.02) |
| Shared |  | 0.83 (0.66, 1.04) |  | 1.09 (0.89, 1.33) |
| None (open defecation) |  | ref |  | ref |
| **Community-level variables** |  |  |  |  |
| Safe disposal of child stool (village-level) [3] (b) | 3475 | 0.97 (0.61, 1.54) | 3474 | 1.06 (0.69, 1.65) |
| Improved sanitation facility (village-level) [2] (b) | 3489 | 0.82 (0.64, 1.06) | 3488 | 0.93 (0.72, 1.20) |
| Shared sanitation facility (village-level) (b) | 3489 | 0.91 (0.56, 1.49) | 3488 | 1.03 (0.64, 1.67) |
| OD (village-level) (b) | 3489 | 1.28 (1.00, 1.63) | 3488 | 1.10 (0.85, 1.42) |
| (a) Adjusted for child gender, child age, child illness, maternal age, maternal education, household size, and household wealth index quintile; clustered by village. (b) Adjusted for village-level covariates: % male, mean child age, % with illness, % breastfed, and mean household wealth index quintile. (c) only children >6 months included, per WHO minimum recommended dietary diversity and meal frequencies. [1] Improved sources of drinking water include: piped water into dwelling/yard/plot, public tap or standpipe, tubewell or borehole, protected dug well, protected spring, bottled water, and rainwater. [2] Improved sanitation facilities include: flush/pour flush toilet to a piped sewer system, septic tank or pit latrine, a ventilated improved pit latrine, a pit latrine with slab, and a composting toilet. [3] Safe disposal of children faeces consist of putting or rinsing stool into any sanitation facility; unsafe disposal of children faeces includes putting or rinsing stool into a drain or ditch, throwing it into garbage, burying or leaving it in the open. |  |  |  |  |

## Discussion

We examined household-level nutrition and WASH characteristics and community-level WASH infrastructure on early childhood linear growth in rural Cambodia. Before adjustment, several WASH and nutrition variables at the child, household, and community level appeared to be associated with improved growth outcomes: breastfeeding of the child, presence of soap and water at the handwashing station, household improved source of drinking water, safe disposal of children’s stools, and household improved sanitation facility (compared to those practicing open defecation) were all associated with reduced odds of stunting and/or increased LAZ score. After adjustment for potential confounders, presence of water and soap at a household’s handwashing station at the time of survey was found to be associated (p<0.05) with higher LAZ score (+0.10, 95% CI: 0.03, 0.16), and household use of an improved drinking water source was associated with less stunting compared to households that did not use an improved source of drinking water (aPR 0.81, 95% CI: 0.66, 0.98). These results underscore the potential role of good hygiene – including handwashing with soap and other practices made possible by more reliable water supply at the household level – in promoting optimal growth outcomes among children^13,40–42^. At the community level, high prevalence of shared sanitation facilities –considered as sub-optimal compared to individual household sanitation in international monitoring^43^ – was found to be negatively associated with child LAZ for children under 12 months of age, suggesting that caregiver WASH practices and exposures as possible routes of transmission for younger infants. These findings are consistent with other studies reporting adverse health effects associated with shared sanitation facilities^44^. No other sanitation variables (household’s safe disposal of child stools, household-level sanitation, community-level sanitation) were associated with LAZ scores or stunting in children under 24 months of age in this study. Our results are consistent with other observational studies reporting associations between WASH and reduced child undernutrition^26,27,45–49^, though such associations have not generally been realised in experimental trials^50,51^. Breastfeeding was associated with reduced length (LAZ −0.16, 95% CI: - 0.27, −0.05); however, other studies have observed that mothers may breastfeed longer if the child is smaller and wean early if the child is physically large^52^. No other measure of feeding practices (dietary diversity, meal frequency, minimum acceptable diet) was associated with growth outcomes in this study.

The most recent CDHS dataset from 2014 (data collection between June-November 2014) reported a mean LAZ of −1.10 (SD 1.52) and 26% (SD 44%) of children stunted among children under 24 months the same provinces (Pursat, Battambang, and Siem Reap), suggesting greater growth faltering in previous surveys compared with ours. These estimates are consistent with the trend of rapidly improving child growth that rural Cambodia has been experiencing in the past 20 years as indicated in CDHS data. While limited to rural communities in three of thirteen provinces of Cambodia, our findings are also consistent with CDHS findings of patterns of preferred sanitation facilities: Cambodian families prefer to move directly from open defecation to “improved” sanitation facilities (pour-flush, with a cleanable slab) rather than incrementally moving up the sanitation ladder (i.e., traditional pit latrines)^53^.

Though the critical window for interventions to increase child linear growth is in the first two years of life, most studies measuring the prevalence of stunting and linear growth have examined older children, typically under 5 years of age. In older children, growth deficits have generally shown a stronger apparent correlation with WASH characteristics in observational studies across geographies. Studies from Peru and Indonesia among children under two and three years of age, respectively, found household sanitation to be associated with taller children^46,47^. Similarly, a meta-analysis that captured data from 70 low- and middle-income countries found household access to an improved sanitation facility to be associated with lower risk of stunting (Odds Ratio of 0.92)^49^ among children under five years of age. In Cambodia, previous observational studies reported strong associations between nutrition and WASH variables on child linear growth and stunting for children. Consistent with our findings, one study using pooled CDHS data from 2000-2005 found no association between feeding indicators (dietary diversity and meal frequency) and child growth outcomes in children aged 6-23 months in Cambodia^54^. Another study using pooled CDHS data from 2000-2010 found household access to an improved sanitation facility to be associated with a lower prevalence of stunting among children under five years (PR 0.82, 95% CI: 0.69-0.96)^27^; the same study performed a subgroup analysis on feeding practices and child growth and did not find any statistically significant associations between exclusive breastfeeding (<6 months) and meal frequency (6-23 months) on stunting. Differences in estimates may be explained by differences in study design and methods, including examining different age strata, variability in measuring risk factors, study setting (e.g., rural versus urban), and timing: Cambodia has experienced rapid growth and development in recent years^55^, with accompanying substantial changes in the prevalence of risk factors that may influence growth outcomes in children.

Observational studies of older children in Ecuador, Mali, and India that have found community-level sanitation to be associated with child growth that may be greater than the effect of household-level sanitation^22,24,56–58^. Similarly, a meta-analysis that included data from 93 countries found that children under five years of age living in communities with high sanitation coverage and no household sanitation facility had lower odds of being stunted than children living in communities with low coverage and with household sanitation, further signalling the role of community^48^. In Cambodia, a previous study of children under five years of age concluded that reduction in children’s exposure to open defecation between 2005-2010 accounted for much or all of the increase in average child height^26^. Such effects may not be discernible in children under 24 months of age but may be apparent in older children as growth trajectories manifest beyond early childhood.

This study adds to a growing body of evidence suggesting that the relationship between water and sanitation infrastructure, hygiene, nutrition, and growth outcomes is complex, variable, and context-specific^51^. Several recent nutrition and WASH trials have been designed and implemented assuming a causal framework linking improved nutrition and WASH to improved child health outcomes, including linear growth and stunting. A systematic review identified five randomised controlled trials that found a small but statistically meaningful effect among children under five years of age^12^; another systematic review of sanitation intervention trials found similar, modest effects of sanitation on nutritional status among children of varying age groups up to school-age (LAZ +0.08, 95% CI: 0.00, 0.16)^59^. The WASH Benefits trials in Kenya and Bangladesh reported growth gains attributable to integrated nutrition and sanitation programming compared to control among children among children under 30 months of age, although these observed gains were likely to have been attributable to nutritional improvements alone since there were no measurable added benefits from adding WASH programming to nutrition^9,10^. Similarly, the SHINE trial in Zimbabwe reported beneficial growth effects among children approximately 18 months of age from nutrition programming but no added benefits of integrating WASH with nutrition programming^21^. Overall, the available evidence for WASH’s role in supporting growth outcomes is mixed, warranting a closer examination of underlying mechanisms driving child growth and a need to expand the scope of transformational WASH interventions that most effectively separate the whole families from faecal exposures.

Our results should be considered alongside the limitations of our methods. The survey data were self-reported and therefore open to recall biases, including courtesy bias (responding in ways perceived to be more pleasing to interviewers), desirability bias (over-reporting of positive perceptions), and acquiescence bias (answering in the affirmative). As a cross-sectional study, we were unable to assess directionality of associations, or infer causality between measured variables. For example, the observed association between growth faltering and ongoing breastfeeding may erroneously implicate breastfeeding as a cause of growth faltering, when it is more probably reflective of a compensatory response to underweight status^52^. Village-scale estimates of coverage may or may not be reflective of a child’s exposure to the environment. Finally, this study only captures exposures at one point in time, but longer-term effects of these exposures may not be apparent until later in life.

Our results suggest that better household access to improved drinking water supplies and handwashing with soap may support improved growth outcomes in early childhood in rural Cambodia. These critical WASH interventions are associated with a range of long-term benefits to health and well-being to families. Hygiene education and interventions to support good hygiene should be integrated with other effective interventions in programs that aim to support maternal and child health where risks of undernutrition are high.

## Data Availability

The data that support the findings of this study are openly available in OSF, DOI 10.17605/OSF.IO/56HPJ.

## Other Information

### Protocols

This study and protocols for data collection and analysis were approved by the National Ethics Committee for Health Research in Cambodia (ref: 110NECHR) and the Internal Review Board at the Georgia Institute of Technology.

## References

1. Dewey, K. G. & Begum, K. Long-term consequences of stunting in early life. Maternal & Child Nutrition 7, 5–18 (2011).

2. Victora, C. G. et al. Maternal and child undernutrition: consequences for adult health and human capital. The Lancet 371, 340–357 (2008).

3. Grantham-McGregor, S. et al. Developmental potential in the first 5 years for children in developing countries. Lancet (London, England) 369, 60–70 (2007).

4. WHO Multicentre Growth Reference Study Group. WHO Child Growth Standards: Length/height-for-age, weight-for-age, weight-for-length, weight-for-height and body mass index-for-age. Methods and development. (2006).

5. Prentice, A. M. et al. Critical windows for nutritional interventions against stunting. The American Journal of Clinical Nutrition 97, 911–918 (2013).

6. Stein, A. D. et al. Growth patterns in early childhood and final attained stature: data from five birth cohorts from low-and middle-income countries. American journal of human biology□: the official journal of the Human Biology Council 22, 353–359 (2010).

7. Mertens, A. et al. Causes and consequences of child growth failure in low-and middle-income countries. medRxiv 2020.06.09.20127100 (2020) doi:10.1101/2020.06.09.20127100.

8. Dewey, K. G. & Adu-Afarwuah, S. Systematic review of the efficacy and effectiveness of complementary feeding interventions in developing countries. Maternal & child nutrition 4 Suppl 1, 24–85 (2008).

9. Null, C. et al. Effects of water quality, sanitation, handwashing, and nutritional interventions on diarrhoea and child growth in rural Kenya: a cluster-randomised controlled trial. The Lancet Global Health 6, e316–e329 (2018).

10. Luby, S. P. et al. Effects of water quality, sanitation, handwashing, and nutritional interventions on diarrhoea and child growth in rural Bangladesh: a cluster randomised controlled trial. The Lancet Global Health 6, e302–e315 (2018).

11. Ruel, M. T. & Alderman, H. Nutrition-sensitive interventions and programmes: how can they help to accelerate progress in improving maternal and child nutrition? Lancet (London, England) 382, 536–551 (2013).

12. Dangour, A. D. et al. Interventions to improve water quality and supply, sanitation and hygiene practices, and their effects on the nutritional status of children. Cochrane Database of Systematic Reviews CD009382 (2013) doi:10.1002/14651858.CD009382.pub2.

13. Mbuya, M. N. N. & Humphrey, J. H. Preventing environmental enteric dysfunction through improved water, sanitation and hygiene: an opportunity for stunting reduction in developing countries. Maternal & Child Nutrition 12, 106–120 (2016).

14. Ngure, F. M. et al. Water, sanitation, and hygiene (WASH), environmental enteropathy, nutrition, and early child development: making the links. Annals of the New York Academy of Sciences 1308, 118–128 (2014).

15. United Nations. SDG Goal 2: End hunger, achieve food security and improved nutrition and promote sustainable agriculture. (2017).

16. National Institute of Statistics, Directorate General for Health, and I. I. Cambodia Demographic and Health Survey 2014.

17. Menon, P., Ruel, M. T. & Morris, S. S. Socioeconomic differentials in child stunting are consistently larger in urban than in rural areas. (2000).

18. Smith, L. C., Ruel, M. T. & Ndiaye, A. Why Is Child Malnutrition Lower in Urban Than in Rural Areas? Evidence from 36 Developing Countries. World Development 33, 1285–1305 (2005).

19. Pickering, A. J., Djebbari, H., Lopez, C., Coulibaly, M. & Alzua, M. L. Effect of a community-led sanitation intervention on child diarrhoea and child growth in rural Mali: A cluster-randomised controlled trial. The Lancet Global Health 3, e701–e711 (2015).

20. Patil, S. R. et al. The Effect of India’s Total Sanitation Campaign on Defecation Behaviors and Child Health in Rural Madhya Pradesh: A Cluster Randomized Controlled Trial. PLOS Medicine 11, e1001709 (2014).

21. Humphrey, J. H. et al. Independent and combined effects of improved water, sanitation, and hygiene, and improved complementary feeding, on child stunting and anaemia in rural Zimbabwe: a cluster-randomised trial. The Lancet Global Health 7, e132–e147 (2019).

22. Fuller, J. A., Villamor, E., Cevallos, W., Trostle, J. & Eisenberg, J. N. I get height with a little help from my friends: herd protection from sanitation on child growth in rural Ecuador. International journal of epidemiology 45, 460–469 (2016).

23. Fuller, J. A. & Eisenberg, J. N. S. Herd Protection from Drinking Water, Sanitation, and Hygiene Interventions. The American journal of tropical medicine and hygiene 95, 1201–1210 (2016).

24. Harris, M., Alzua, M. L., Osbert, N. & Pickering, A. Community-Level Sanitation Coverage More Strongly Associated with Child Growth and Household Drinking Water Quality than Access to a Private Toilet in Rural Mali. Environmental Science & Technology 51, 7219–7227 (2017).

25. Spears, D. Exposure to open defecation can account for the Indian enigma of child height. Journal of Development Economics 146, 102277 (2020).

26. Vyas, S., Kov, P., Smets, S. & Spears, D. Disease externalities and net nutrition: Evidence from changes in sanitation and child height in Cambodia, 2005–2010. Economics & Human Biology 23, 235–245 (2016).

27. Ikeda, N., Irie, Y. & Shibuya, K. Determinants of reduced child stunting in Cambodia: analysis of pooled data from three demographic and health surveys. Bulletin of the World Health Organization 91, 341–349 (2013).

28. Prendergast, A. J. et al. Independent and combined effects of improved water, sanitation, and hygiene, and improved complementary feeding, on stunting and anaemia among HIV-exposed children in rural Zimbabwe: a cluster-randomised controlled trial. The Lancet Child & Adolescent Health 3, 77–90 (2019).

29. Briceño, B., Coville, A., Gertler, P. & Martinez, S. Are there synergies from combining hygiene and sanitation promotion campaigns: Evidence from a large-scale cluster-randomized trial in rural Tanzania. PloS one 12, e0186228–e0186228 (2017).

30. Cameron, L., Olivia, S. & Shah, M. Scaling up sanitation: Evidence from an RCT in Indonesia. Journal of Development Economics 138, 1–16 (2019).

31. Clasen, T. et al. Effectiveness of a rural sanitation programme on diarrhoea, soil-transmitted helminth infection, and child malnutrition in Odisha, India: a cluster-randomised trial. The Lancet Global Health 2, e645–e653 (2014).

32. Cogill, B. Anthropometric Indicators Measurement Guide. (2001).

33. Faraone, S. V. Interpreting estimates of treatment effects: implications for managed care. P & T□: a peer-reviewed journal for formulary management 33, 700–711 (2008).

34. Barros, A. J. D. & Hirakata, V. N. Alternatives for logistic regression in cross-sectional studies: an empirical comparison of models that directly estimate the prevalence ratio. BMC Medical Research Methodology 3, 21 (2003).

35. Thompson, C. G., Kim, R. S., Aloe, A. M. & Becker, B. J. Extracting the Variance Inflation Factor and Other Multicollinearity Diagnostics from Typical Regression Results. Basic and Applied Social Psychology 39, 81–90 (2017).

36. VanderWeele, T. J. Principles of confounder selection. European Journal of Epidemiology 34, 211–219 (2019).

37. World Health Organization. Indicators for assessing infant and young child feeding practices. Part I: Definitions. (2008).

38. WHO/UNICEF JMP for Water Supply and Sanitation. WASH Post-2015: Proposed indicators for drinking water, sanitation and hygiene. (2015).

39. Bauza, V., Reese, H., Routray, P. & Clasen, T. Child Defecation and Faeces Disposal Practices and Determinants among Households after a Combined Household-Level Piped Water and Sanitation Intervention in Rural Odisha, India. The American Journal of Tropical Medicine and Hygiene 100, 1013–1021 (2019).

40. von Salmuth, V. et al. Maternal-focused interventions to improve infant growth and nutritional status in low-middle income countries: A systematic review of reviews. PloS one 16, e0256188 (2021).

41. Black, R. E. Would control of childhood infectious diseases reduce malnutrition? Acta paediatrica Scandinavica. Supplement 374, 133–140 (1991).

42. Langford, R., Lunn, P. & Panter-Brick, C. Hand-washing, subclinical infections, and growth: a longitudinal evaluation of an intervention in Nepali slums. American journal of human biology□: the official journal of the Human Biology Council 23, 621–629 (2011).

43. World Health Organization (WHO) and the United Nations Children’s Fund (UNICEF). Progress on household drinking water, sanitation and hygiene 2000-2020: five years into the SDGs. https://washdata.org/sites/default/files/2021-07/jmp-2021-wash-households.pdf (2021).

44. Heijnen, M. et al. Shared Sanitation versus Individual Household Latrines: A Systematic Review of Health Outcomes. PLOS ONE 9, e93300 (2014).

45. Spears, D. How Much International Variation in Child Height Can Sanitation Explain? World Bank Policy Research Working Paper No. 6351 (2013).

46. Checkley, W. et al. Effect of water and sanitation on childhood health in a poor Peruvian peri-urban community. The Lancet 363, 112–118 (2004).

47. Rah, J. H., Sukotjo, S., Badgaiyan, N., Cronin, A. A. & Torlesse, H. Improved sanitation is associated with reduced child stunting amongst Indonesian children under 3 years of age. Maternal & Child Nutrition 16, e12741 (2020).

48. Larsen, D. A., Grisham, T., Slawsky, E. & Narine, L. An individual-level meta-analysis assessing the impact of community-level sanitation access on child stunting, anemia, and diarrhea: Evidence from DHS and MICS surveys. PLOS Neglected Tropical Diseases 11, e0005591 (2017).

49. Fink, G., Günther, I. & Hill, K. The effect of water and sanitation on child health: evidence from the demographic and health surveys 1986–2007. International Journal of Epidemiology 40, 1196–1204 (2011).

50. Cumming, O. et al. The implications of three major new trials for the effect of water, sanitation and hygiene on childhood diarrhea and stunting: a consensus statement. BMC Medicine 17, 173 (2019).

51. Rogawski McQuade, E. T., Benjamin-Chung, J., Westreich, D. & Arnold, B. F. Population intervention effects in observational studies to emulate target trial results: reconciling the effects of improved sanitation on child growth. International Journal of Epidemiology (2021) doi:10.1093/ije/dyab070.

52. Marquis, G. S., Habicht, J. P., Lanata, C. F., Black, R. E. & Rasmussen, K. M. Association of breastfeeding and stunting in Peruvian toddlers: an example of reverse causality. International journal of epidemiology 26, 349–356 (1997).

53. United Nations Children’s Fund (UNICEF) and World Health Organization. Progress on household drinking water, sanitation and hygiene 2000-2017. Special focus on inequalities. (2019).

54. Marriott, B. P., White, A. J., Hadden, L., Davies, J. C. & Wallingford, J. C. How well are infant and young child World Health Organization (WHO) feeding indicators associated with growth outcomes? An example from Cambodia. Maternal & Child Nutrition 6, 358–373 (2010).

55. World Bank. World Development Indicators: Economic Policy & Debt Aggregate Indicators. https://data.worldbank.org/indicator.

56. Kov, P., Smets, S., Spears, D. & Vyas, S. Growing taller among toilets: Evidence from changes in sanitation and child height in Cambodia, 2005-2010. (2013).

57. Hammer, J. & Spears, D. Village sanitation and child health: Effects and external validity in a randomized field experiment in rural India. Journal of Health Economics 48, 135–148 (2016).

58. Spears, D. Exposure to open defecation can account for the Indian enigma of child height. Journal of Development Economics 146, 102277 (2020).

59. Freeman, M. C. et al. The impact of sanitation on infectious disease and nutritional status: A systematic review and meta-analysis. International Journal of Hygiene and Environmental Health 220, 928–949 (2017).

