## Supplemental Information for "Risk factors for early childhood growth faltering in rural Cambodia"

### Abstract

#### Introduction

Inadequate nutrition in early life and exposure to sanitation-related enteric pathogens have been linked to poor growth outcomes in children. Despite rapid development in Cambodia, high prevalence of growth faltering and stunting continue to persist. This study aimed to assess nutrition and WASH variables and their association with nutritional status of children under 24 months in rural Cambodia.

#### Methods

We conducted surveys in 491 villages across 55 rural communes in Cambodia in September 2016 to measure associations between child, household, and community-level risk factors for stunting and length-for-age z-score (LAZ). A primary survey measured child-level variables, including anthropometric measures and risk factors for growth faltering and stunting, for 4,036 children under 24 months of age from 3,877 households (approximately 8 households per village). For LAZ, we calculated bivariate and adjusted associations (as mean differences) with 95% confidence intervals using generalised estimating equations (GEEs) to fit linear regression models with robust standard errors. For stunting, we calculated unadjusted and adjusted prevalence ratios (PRs) with 95% confidence intervals using GEEs to fit Poisson regression models with robust standard errors. For all models assessing effects of household-level variables, we used GEEs to account for clustering at the village level.

#### Results

After adjustment for potential confounding, presence of water and soap at a household’s handwashing station was found to be significantly associated (p<0.05) with increased LAZ (adjusted mean difference in LAZ +0.10, 95% CI: 0.03, 0.16), and household use of an improved drinking water source was associated with less stunting in children compared to households that did not use an improved source of drinking water (aPR 0.81, 95% CI: 0.66, 0.98); breastfeeding was associated with a lower LAZ score (-0.16, 95% CI: -0.27, -0.05). No other feeding practices (i.e., dietary diversity, meal frequency, minimum acceptable diet) or sanitation variables (i.e., household’s safe disposal of child stools, household-level sanitation, community-level sanitation) were associated with LAZ scores or stunting in children under 24 months of age. In an age-stratified analysis, children under 12 months of age were longer (LAZ +0.12, 95% CI: 0.02, 0.21) if there was presence of water and soap at the household handwashing station; at the community level, higher prevalence of shared sanitation (percentage of households in a village who report to use shared sanitation facilities) was negatively associated with child length (LAZ -0.36, 95% CI: -0.66, -0.07).

### Supplemental Information

#### Survey design

All surveys were communicated in the Khmer language, spoken as a first language by 100% of residents in the study area^1^. For each of the 491 villages, 8 households with at least one child under 24 months were randomly selected for the primary household survey to assess the eligible children’s diet and nutritional status. This survey was administered to the mother or other primary caregiver (91% of respondents were mothers; primary caregiver was surveyed when the mother was unavailable). The primary survey included socioeconomic and demographic questions, including child age and sex; household assets; caregiver education level; breastfeeding and nutrition practices; and sanitation and hygiene behaviours. We calculated household wealth using an asset-based wealth index using methodology provided by the CDHS^2^, constructed using principal component analysis (PCA) and excluding WASH-variables in order to evaluate associations between wealth and sanitation coverage. The primary survey also included infant and young child feeding (IYCF) indicators suggested by the WHO include minimum dietary diversity, minimum meal frequency, and minimum adequate diet^3–5^ for infants and children 6-23 months. WHO dietary diversity score consists of categorizing solid foods into eight food groups^6^, including: breastmilk, grains, legumes/nuts, dairy, flesh meat, eggs, vitamin-A-rich fruits and vegetables, and other fruits and vegetables. To suit the Cambodian context, the evaluation team asked additional questions on the types of fish and other wild animals consumed, which are included in the flesh meat group. The dietary diversity score is on a scale from 0 – 8 and determined based on the number of food groups the caregiver reported to have fed the child in the last 24 hours; minimum dietary diversity is defined as having received food from five or more food groups (or a dietary diversity score greater than or equal to five). Minimum meal frequency is defined by the frequency of solid and semi-solid foods received based on a child’s age and whether the child is breastfed. The minimum number of times breastfed children should receive solid, semi-solid, or soft foods varies with age (2 times if 6–8 months and 3 times if 9–23 months). The minimum number of times non-breastfed children should receive solid, semi-solid, or soft foods, including milk, is 4 times for all children 6–23 months (does not vary by age). Sanitation facilities were observed and verified by enumerators and recorded based on CDHS methodology.

We conducted a secondary survey to record information on community-level WASH variables. This included questions on household WASH conditions among the 8 selected households with children under 24 months and an additional 3 households randomly selected among all households in the same areas. The secondary 10-minute survey and structured observation was conducted to assess WASH characteristics regardless of whether there were children in the household to determine whether village-level mean and variance of WASH-variables scores were associated with improved child health outcomes. Enumerators visually observed and recorded WASH conditions in the household and took photographs of household latrines, which were tagged with the unique household identification number for verification of proper classification. A random sample of 20% of the photos were cross-checked with the recorded survey to ensure proper classification. Together, at the village level, there were 8 households with key outcome measurements and 11 households with key exposures of WASH at the community level. Given the oversampling of households with children under 24 months of age, post-stratification weights were used to get a representative sample of the population. Sampling weights calculated as follows: first, we estimated the proportion of households with children under age 2 at the village-level based on conversations with village leaders. This estimate was then divided by the proportion of sampled households with children under 24 months of age at each village to yield the sampling weight for each household from the main sample. For the three additional households, the sampling weights were calculated by dividing the remaining proportion of total households at the village level by the proportion of sampled households at each village. This results in underweighting the households with children under 24 months of age and overweighting the supplemental households.

#### Sample size and power calculations.

The sample size calculations are anchored to a future randomized control trial and powered to detect an MDES of 0.18 LAZ, similar to other trials^7^.

The following sample size calculations for the primary outcome, difference in mean LAZ scores between treatment groups, were conducted based on different ICC scenarios. This is revised to account for a drop of three treatment communes after randomization occurred. Power calculations assume α=0.05, power=0.8, mean baseline LAZ estimate of -1.637 with a standard deviation of 1.286, and a two-sided test for a two-sample comparison of means. LAZ calculations use a standard equation assuming a single, post-treatment measurement at 2 years.

| MDES between treatment groups | Subjects per cluster (commune) | Estimated total number of subjects required (all 4 arms) |
| --- | --- | --- |
| ICC=0.01 | | |
| 0.15 | 155 | 8,525 |
| 0.16 | 115 | 6,325 |
| 0.17 | 90 | 4,950 |
| **0.18** | **73** | **4,015** |
| 0.19 | 61 | 3,355 |
| 0.2 | 52 | 2,860 |
| ICC=0.015 | | |
| 0.15 | 690 | 37,950 |
| 0.16 | 268 | 14,740 |
| 0.17 | 162 | 8,910 |
| 0.18 | 114 | 6,270 |
| 0.19 | 87 | 4,785 |
| 0.2 | 70 | 3,850 |
| ICC=0.02 | | |
| 0.15 | NA | NA |
| 0.16 | NA | NA |
| 0.17 | 876 | 48,180 |
| 0.18 | 268 | 14,740 |
| 0.19 | 155 | 8,525 |
| 0.2 | 107 | 5,885 |

For the main sample, in order to collect reliable point-estimates of child growth at the group level, the required sample size was calculated based on a conventional approach for means at the 95 percent confidence level with a margin of error of +/-6 percent.

$$N=D_{eff}*\left[ \frac{\left( Z*\sigma\right)}{ME} \right]^{2}*4=1.4*\left[ \frac{\left( 1.96*1.286 \right)}{0.098} \right]^{2}*4=3,705$$

where:

$D_{eff}=$design effect of 1.4, estimated using 2014 DHS data

$Z=$ 1.96 (for 95% confidence level)

$\sigma=$ child growth standard deviation of 1.286, estimated using DHS 2014 data

$ME=$ 0.098, corresponding to margin of error of +/-6%

Dividing the minimum required sample of 3,705 by 491 villages results in 7.54 surveys per village. To ease logistics and provide a cushion for unexpected challenges, sample size was rounded up to 8 surveys per village, resulting in a target total of 3,928 main households.

For the secondary sample, in order to collect reliable point-estimates of sanitation coverage at the group level, the required sample size was calculated based on a conventional approach for proportions, at the 95 percent confidence level with a margin of error of +/-5 percent.

$$N=p(1-p)*\left[ \frac{Z}{ME} \right]^{2}*4=0.408(1-0.592)*\left[ \frac{1.96}{0.05} \right]^{2}*4=1,024$$

where:

$p=$proportion of sanitation coverage of 0.408, estimated using DHS 2014 data

$Z=$ 1.96 (for 95% confidence level)

$ME=$ margin of error of +/-5%

Given the 491 villages, sample size was rounded up to three additional randomly selected households per village, for a target total of 1,473 additional households, as shown below.

Required Sample Size

| **Provinces** | **Communes** | **Villages** | **HHs Main Sample** | **HHs Secondary Sample** |
| --- | --- | --- | --- | --- |
| Battambang | 22 | 180 | 1,440 | 540 |
| Pursat | 6 | 83 | 664 | 249 |
| Siem Reap | 27 | 228 | 1,824 | 684 |
| **Total** | **55** | **491** | **3,928** | **1,473** |

#### Anthropometry protocols

Anthropometric measurement is comprised of weight and length. Weight was measured using Uniscale (UNICEF recommended scale) in Kilogram with precision to one decimal point. Length was measured using a length board (UNICEF / WFP recommended) in Centimetre with precision to one decimal point. Two data measurements were required, one from the measurement taker and another one from an assistant. The measurement procedure followed FANTA Guidelines:

- Weight measurement:
- *Preparation*: Ensure enough material is available for measurement (scale, battery, pen, tissue, record form, and age calculation form) with proper function. Ensure that the scale is positioned in a plate and smooth surface. Measurement taker is on the right hand of mother/caregiver while assistant is in front of mother/caregiver. Ensure that children dress light clothes with no cap or shoes. Assistant helps mother/caregiver in carrying the child and asks mother/caregiver to go on to the scale after proper functioning.
- *During Measurement*: Request mother/caregiver to stand on the scale, inform the measurement result loudly, press button to measure child, hand the child to mother/caregiver after scale functioning, read weight of child out loud so that assistant can record the measurement.
- *Second Measurement*: Request mother/caregiver to step off the scale. Repeat the measurement steps. Record second measurement.
- Length measurement:
- *Preparation*: Prepare length board on a plate and smooth surface. Ensure length board stability, take off shoe and cap from child. Check measurement level on the length board, and ensure the record form.
- *During Measurement*: Lay child on his back on the board, check head, eye, shoulder, hand, buttock, knees, and heel. Make sure body is in proper position and still. Measurement must be read to the nearest of 0.1 cm. Repeat the measurement one more time to ensure accuracy of reading. If the two measurements are different by more than 1.0 cm, then a third measurement is taken.
- Following the weighing and length measurements, any child who is classified as severely malnourished is referred to the health clinic.
- Training: The enumerators were trained on the protocols to follow and how to calibrate equipment. Tested on accurate recording of length measurements. Hands-on practice in pairs and then we did a standardization exercise where the entire team is tested on their ability to measure child length accurately and precisely. Measurers had to meet the accuracy and precision threshold to pass and be hired as enumerators for data collection.
- Field supervision: The anthropometry specialist was present in the field during the entire baseline phase, accompanying enumerators to ensure proper technique with the height and weight measurements and recording.

#### Table: Variables associated with village-mean LAZ

|  | Bivariate (unadjusted) | | | | Adjusted | | | |
| --- | --- | --- | --- | --- | --- | --- | --- | --- |
|  | LAZ effect | 95% CI | n |  | LAZ effect | 95% CI | n |  |
| Mean child birth weight (kg) | 0.84 | (0.63, 1.05) | 491 | *** | 0.79 | (0.59, 0.99) | 491 | *** |
| % minimum dietary diversity met | 0.13 | (-0.05, 0.31) | 491 |  | -0.07 | (-0.26, 0.11) | 491 |  |
| % minimum meal frequency met | -0.22 | (-0.42, -0.02) | 491 | ** | 0.07 | (-0.16, 0.29) | 491 |  |
| % improved drinking water source [1] | 0.04 | (-0.17, 0.24) | 491 |  | -0.04 | (-0.23, 0.16) | 491 |  |
| % improved sanitation facility [2] | 0.15 | (0.00, 0.30) | 491 | ** | 0.01 | (-0.16, 0.18) | 491 |  |
| % safe disposal of child stool [3] | -0.05 | (-0.25, 0.15) | 488 |  | 0.05 | (-0.14, 0.24) | 488 |  |
| % OD | -0.12 | (-0.26, 0.01) | 491 |  | 0.02 | (-0.13, 0.17) | 491 |  |
| % with shared sanitation facility | 0.02 | (-0.18, 0.22) | 491 |  | -0.05 | (-0.24, 0.14) | 491 |  |
| % with finished floor | -0.14 | (-0.57, 0.30) | 491 |  | -0.08 | (-0.50, 0.33) | 491 |  |
| % male | -0.11 | (-0.32, 0.09) | 491 |  | -- |  | -- |  |
| Mean child age (months) | -0.05 | (-0.06, -0.03) | 491 | *** | -- |  | -- |  |
| Mean HH wealth index quintile (non-WASH) | 0.09 | (0.04, 0.13) | 491 | *** | -- |  | -- |  |
| % with any illness [4] | -0.04 | (-0.24, 0.16) | 491 |  | -- |  | -- |  |
| % currently breastfed | 0.41 | (0.18, 0.65) | 491 | *** | -- |  | -- |  |
| **Village-level WASH variables (using primary and secondary surveys with post-stratification weights)** | | | | | | | | |
| % improved drinking water source [1] | -0.16 | (-0.29, -0.03) | 491 | ** | -0.15 | (-0.27, -0.02) | 491 | ** |
| % improved sanitation facility [2] | 0.13 | (0.01, 0.25) | 491 | ** | 0.06 | (-0.07, 0.18) | 491 |  |
| % safe disposal of child stool [3] | -0.06 | (-0.30, 0.17) | 488 |  | -0.08 | (-0.30, 0.14) | 488 |  |
| % OD | -0.09 | (-0.21, 0.03) | 491 |  | 0.01 | (-0.12, 0.14) | 491 |  |
| % with shared sanitation facility | -0.13 | (-0.37, 0.11) | 491 |  | -0.20 | (-0.43, 0.02) | 491 |  |
| **p<0.05; ***p<0.01; Adjusted analyses are adjusted for the following village-level variables: child gender, child age, household wealth, child illness, and breastfeeding status. [1] Improved sources of drinking water include: piped water into dwelling/yard/plot, public tap or standpipe, tube well or borehole, protected dug well, protected spring, bottled water, and rainwater. [2] Improved sanitation facilities include: flush/pour flush toilet to a piped sewer system, septic tank or pit latrine, a ventilated improved pit latrine, a pit latrine with slab, and a composting toilet. [3] Safe disposal of children faeces consist of putting or rinsing stool into a sanitation facility or burying it; unsafe disposal of children faeces includes putting or rinsing stool into a drain or ditch, throwing it into garbage or leaving it in the open. [4] Any child illness includes 7-day recall of caregiver-reported vomiting, abdominal pain, blood in stool, or diarrhoea (7 and 14-day recall) | | | | | | | | |

#### Table: Variables associated with village-level stunting

|  | Bivariate (unadjusted) | | | | Adjusted | | | |
| --- | --- | --- | --- | --- | --- | --- | --- | --- |
|  | OR | 95% CI | n |  | OR | 95% CI | n |  |
| Mean child birth weight (kg) | 0.84 | (0.63, 1.05) | 491 | *** | 0.79 | (0.59, 0.99) | 491 | *** |
| % minimum dietary diversity met | 0.13 | (-0.05, 0.31) | 491 |  | -0.07 | (-0.26, 0.11) | 491 |  |
| % minimum meal frequency met | -0.22 | (-0.42, -0.02) | 491 | ** | 0.07 | (-0.16, 0.29) | 491 |  |
| % improved drinking water source [1] | 0.04 | (-0.17, 0.24) | 491 |  | -0.04 | (-0.23, 0.16) | 491 |  |
| % improved sanitation facility [2] | 0.15 | (0.00, 0.30) | 491 | ** | 0.01 | (-0.16, 0.18) | 491 |  |
| % safe disposal of child stool [3] | -0.05 | (-0.25, 0.15) | 488 |  | 0.05 | (-0.14, 0.24) | 488 |  |
| % OD | -0.12 | (-0.26, 0.01) | 491 |  | 0.02 | (-0.13, 0.17) | 491 |  |
| % with shared sanitation facility | 0.02 | (-0.18, 0.22) | 491 |  | -0.05 | (-0.24, 0.14) | 491 |  |
| % with finished floor | -0.14 | (-0.57, 0.30) | 491 |  | -0.08 | (-0.50, 0.33) | 491 |  |
| % male | -0.11 | (-0.32, 0.09) | 491 |  | -- |  | -- |  |
| Mean child age (months) | -0.05 | (-0.06, -0.03) | 491 | *** | -- |  | -- |  |
| Mean HH wealth index quintile (non-WASH) | 0.09 | (0.04, 0.13) | 491 | *** | -- |  | -- |  |
| % with any illness [4] | -0.04 | (-0.24, 0.16) | 491 |  | -- |  | -- |  |
| % currently breastfed | 0.41 | (0.18, 0.65) | 491 | *** | -- |  | -- |  |
| **Village-level WASH variables (using primary and secondary surveys with post-stratification weights)** | | | | | | | | |
| % improved drinking water source [1] | -0.16 | (-0.29, -0.03) | 491 | ** | -0.15 | (-0.27, -0.02) | 491 | ** |
| % improved sanitation facility [2] | 0.13 | (0.01, 0.25) | 491 | ** | 0.06 | (-0.07, 0.18) | 491 |  |
| % safe disposal of child stool [3] | -0.06 | (-0.30, 0.17) | 488 |  | -0.08 | (-0.30, 0.14) | 488 |  |
| % OD | -0.09 | (-0.21, 0.03) | 491 |  | 0.01 | (-0.12, 0.14) | 491 |  |
| % with shared sanitation facility | -0.13 | (-0.37, 0.11) | 491 |  | -0.20 | (-0.43, 0.02) | 491 |  |
| **p<0.05; ***p<0.01; Adjusted analyses are adjusted for the following village-level variables: child gender, child age, household wealth, child illness, and breastfeeding status. [1] Improved sources of drinking water include: piped water into dwelling/yard/plot, public tap or standpipe, tube well or borehole, protected dug well, protected spring, bottled water, and rainwater. [2] Improved sanitation facilities include: flush/pour flush toilet to a piped sewer system, septic tank or pit latrine, a ventilated improved pit latrine, a pit latrine with slab, and a composting toilet. [3] Proper disposal of children faeces consist of putting or rinsing stool into a sanitation facility or burying it; unsafe disposal of children faeces includes putting or rinsing stool into a drain or ditch, throwing it into garbage or leaving it in the open. [4] Any child illness includes 7-day recall of caregiver-reported vomiting, abdominal pain, blood in stool, or diarrhoea (7 and 14-day recall) | | | | | | | | |

#### Table: Linear regression coefficient for association between nutrition and WASH variables and linear growth, stratified by age groups (1-12 months and >12-24 months)

|  | N | Adjusted effect size  (0-12 months) | N | Adjusted effect size  (>12-24 months) |
| --- | --- | --- | --- | --- |
| **Child-level indicators** |  |  |  |  |
| Currently breastfed (a) | 2145 | **-0.429 (-0.747, -0.111)** | 1564 | -0.112 (-0.227, 0.004) |
| Minimum dietary diversity met (a,c) | 1112 | -0.007 (-0.145, 0.131) | 1598 | 0.063 (-0.033, 0.159) |
| Minimum meal frequency met (a,c) | 1112 | -0.019 (-0.201, 0.163) | 1598 | 0.009 (-0.117, 0.136) |
| **Household-level indicators** |  |  |  |  |
| Improved drinking water source [1] (a) | 2173 | -0.031 (-0.157, 0.094) | 1594 | 0.134 (-0.008, 0.276) |
| Presence of water and soap at handwashing (a) | 2174 | **0.116 (0.025, 0.207)** | 1597 | 0.064 (-0.035, 0.162) |
| Adequate disposal of child stool [4] (a) | 1507 | 0.020 (-0.110, 0.150) | 1336 | 0.095 (-0.128, 0.318) |
| Sanitation facility (a) | 2177 |  | 1592 |  |
| Improved [3] |  | 0.085 (-0.028, 0.198) |  | 0.043 (-0.074, 0.159) |
| Shared |  | -0.029 (-0.157, 0.099) |  | 0.037 (-0.127, 0.201) |
| None (open defecation) |  | ref |  | ref |
| **Community-level indicators** |  |  |  |  |
| Improved drinking water source (village-level) [1] (b) | 2188 | **-0.250 (-0.416, -0.084)** | 1604 | -0.067 (-0.247, 0.112) |
| Adequate disposal of child stool (village-level) [4] (b) | 2179 | -0.306 (-0.621, 0.009) | 1599 | 0.226 (-0.096, 0.548) |
| Improved sanitation facility (village-level) [3] (b) | 2188 | 0.106 (-0.057, 0.268) | 1604 | 0.077 (-0.102, 0.256) |
| Shared sanitation facility (village-level) (b) | 2188 | **-0.362 (-0.657, -0.068)** | 1604 | 0.037 (-0.266, 0.339) |
| OD (village-level) (b) | 2188 | 0.004 (-0.163, 0.170) | 1604 | -0.116 (-0.297, 0.066) |
| (a) Adjusted for child gender, child age, child illness, maternal age, maternal education, household size, and household wealth index quintile; clustered by village. (b) Adjusted for village-level covariates: % male, mean child age, % with illness, % breastfed, and mean household wealth index quintile. (c) only children >6 months included because minimum recommended dietary diversity and meal frequencies. [1] Improved sources of drinking water include: piped water into dwelling/yard/plot, public tap or standpipe, tubewell or borehole, protected dug well, protected spring, bottled water, and rainwater. [2] Improved sanitation facilities include: flush/pour flush toilet to a piped sewer system, septic tank or pit latrine, a ventilated improved pit latrine, a pit latrine with slab, and a composting toilet. [3] Proper disposal of children faeces consist of putting or rinsing stool into a sanitation facility or burying it; inadequate disposal of children faeces includes putting or rinsing stool into a drain or ditch, throwing it into garbage or leaving it in the open. [4] Any child illness includes 7-day recall of caregiver-reported vomiting, abdominal pain, blood in stool, or diarrhoea (7 and 14-day recall) | | | | |

#### Table: STROBE Statement—Checklist of items that should be included in reports of ***cross-sectional studies***

|  | Item No | Recommendation | Page No |
| --- | --- | --- | --- |
| **Title and abstract** | 1 | (*a*) Indicate the study’s design with a commonly used term in the title or the abstract | X |
|  |  | (*b*) Provide in the abstract an informative and balanced summary of what was done and what was found | X |
| Introduction | | | |
| Background/rationale | 2 | Explain the scientific background and rationale for the investigation being reported | X |
| Objectives | 3 | State specific objectives, including any prespecified hypotheses | X |
| Methods | | | |
| Study design | 4 | Present key elements of study design early in the paper | X |
| Setting | 5 | Describe the setting, locations, and relevant dates, including periods of recruitment, exposure, follow-up, and data collection | X |
| Participants | 6 | (*a*) Give the eligibility criteria, and the sources and methods of selection of participants | X |
| Variables | 7 | Clearly define all outcomes, exposures, predictors, potential confounders, and effect modifiers. Give diagnostic criteria, if applicable | X |
| Data sources/ measurement | 8* | For each variable of interest, give sources of data and details of methods of assessment (measurement). Describe comparability of assessment methods if there is more than one group | X |
| Bias | 9 | Describe any efforts to address potential sources of bias | X |
| Study size | 10 | Explain how the study size was arrived at | X |
| Quantitative variables | 11 | Explain how quantitative variables were handled in the analyses. If applicable, describe which groupings were chosen and why | X |
| Statistical methods | 12 | (*a*) Describe all statistical methods, including those used to control for confounding | X |
|  |  | (*b*) Describe any methods used to examine subgroups and interactions | X |
|  |  | (*c*) Explain how missing data were addressed | N/A |
|  |  | (*d*) If applicable, describe analytical methods taking account of sampling strategy | X |
|  |  | (*e*) Describe any sensitivity analyses | N/A |
| Results | | | |
| Participants | 13* | (a) Report numbers of individuals at each stage of study—eg numbers potentially eligible, examined for eligibility, confirmed eligible, included in the study, completing follow-up, and analysed | X |
|  |  | (b) Give reasons for non-participation at each stage | X |
|  |  | (c) Consider use of a flow diagram | X |
| Descriptive data | 14* | (a) Give characteristics of study participants (eg demographic, clinical, social) and information on exposures and potential confounders | X |
|  |  | (b) Indicate number of participants with missing data for each variable of interest | X |
| Outcome data | 15* | Report numbers of outcome events or summary measures | X |
| Main results | 16 | (*a*) Give unadjusted estimates and, if applicable, confounder-adjusted estimates and their precision (eg, 95% confidence interval). Make clear which confounders were adjusted for and why they were included | X |
|  |  | (*b*) Report category boundaries when continuous variables were categorized | X |
|  |  | (*c*) If relevant, consider translating estimates of relative risk into absolute risk for a meaningful time period | X |
| Other analyses | 17 | Report other analyses done—eg analyses of subgroups and interactions, and sensitivity analyses | X |
| Discussion | | | |
| Key results | 18 | Summarise key results with reference to study objectives | X |
| Limitations | 19 | Discuss limitations of the study, taking into account sources of potential bias or imprecision. Discuss both direction and magnitude of any potential bias | X |
| Interpretation | 20 | Give a cautious overall interpretation of results considering objectives, limitations, multiplicity of analyses, results from similar studies, and other relevant evidence | X |
| Generalisability | 21 | Discuss the generalisability (external validity) of the study results | X |
| Other information | | | |
| Funding | 22 | Give the source of funding and the role of the funders for the present study and, if applicable, for the original study on which the present article is based | X |

*Give information separately for exposed and unexposed groups.

**Note:** An Explanation and Elaboration article discusses each checklist item and gives methodological background and published examples of transparent reporting. The STROBE checklist is best used in conjunction with this article (freely available on the Web sites of PLoS Medicine at http://www.plosmedicine.org/, Annals of Internal Medicine at http://www.annals.org/, and Epidemiology at http://www.epidem.com/). Information on the STROBE Initiative is available at www.strobe-statement.org.

#### References

1.	National Institute of Statistics, Directorate General for Health, and I. I. *Cambodia Demographic and Health Survey 2014*.

2.	Rustein, S. O. *Steps to constructing the new DHS wealth index*. (2015).

3.	World Health Organization. Indicators for assessing infant and young child feeding practicies. Part I: Definitions. (2008).

4.	Heidkamp, R. A. *et al.* Implications of Updating the Minimum Dietary Diversity for Children Indicator for Tracking Progress in the Eastern and Southern Africa Region. *Current Developments in Nutrition* **4**, (2020).

5.	Cheung, S. T. *et al.* Albumin mRNA in plasma predicts post-transplant recurrence of patients with hepatocellular carcinoma. *Transplantation* **85**, 81–87 (2008).

6.	Cheung, S. T. *et al.* Albumin mRNA in plasma predicts post-transplant recurrence of patients with hepatocellular carcinoma. *Transplantation* **85**, 81–87 (2008).

7.	Pickering, A. J., Djebbari, H., Lopez, C., Coulibaly, M. & Alzua, M. L. Effect of a community-led sanitation intervention on child diarrhoea and child growth in rural Mali: A cluster-randomised controlled trial. *The Lancet Global Health* **3**, e701–e711 (2015).
